# Democratizing Scientific Publishing: A Local, Multi-Agent LLM Framework for Objective Manuscript Editing

**DOI:** 10.64898/2026.04.13.26350761

**Authors:** Rohan Bhansali, Alon Gorenshtein, Brandon Westover, Daniel M. Goldenholz

## Abstract

Manuscript preparation is a critical bottleneck in scientific publishing, yet existing AI writing tools require cloud transmission of sensitive content, creating data-confidentiality barriers for clinical researchers. We introduce the Paper Analysis Tool (PAT), a free, multi-agent framework that deploys 31 specialized agents powered by small language models (SLMs) to audit manuscripts across multiple quality dimensions without external data transmission. Applied to three published clinical neurological papers, PAT generated 540 evaluable suggestions. Validation by two expert reviewers (R.B., A.G.) confirmed 391 actionable, high-value revisions (90% agreement), achieving a 72.4% overall usefulness accuracy spanning methodological, statistical, and visual domains. Furthermore, deterministic re-evaluation of 126 agent-suggested rewrite pairs using Phase 0 metrics confirmed text improvement: total word count decreased by 25%, passive voice prevalence dropped sharply from 35% to 5%, average sentence length decreased by 24%, long-sentence fraction fell by 67%, and the Flesch-Kincaid grade improved by 17% . Our validation confirms that systematic, agent-driven pre-submission review drives measurable improvements, successfully converting manuscript optimization from an opaque, manual endeavor into a transparent and rigorous scientific process.

Manuscript preparation is a critical bottleneck in scientific publishing, yet existing AI writing tools require cloud transmission of sensitive content, creating data-confidentiality barriers for clinical researchers. We introduce the Paper Analysis Tool (PAT), a free, multi-agent framework that deploys 31 specialized agents powered by small language models (SLMs) to audit manuscripts across multiple quality dimensions without external data transmission. Applied to three published clinical neurological papers, PAT generated 540 evaluable suggestions. Independent validation by two expert reviewers (R.B., A.G.) confirmed 391 actionable, high-value revisions (90% agreement), achieving a 72.4% overall usefulness accuracy spanning methodological, statistical, and visual domains. Furthermore, deterministic re-evaluation of 126 suggested Phase 0 rewrite pairs confirmed text improvement: total word count decreased by 25%, passive voice prevalence dropped sharply from 35% to 5%, average sentence length decreased by 24%, and long-sentence fraction fell by 67%, and the Flesch–Kincaid grade improved modestly. Our validation confirms that systematic, agent-driven pre-submission review drives measurable improvements, successfully converting manuscript optimization from an opaque, manual endeavor into a transparent and rigorous scientific process.

## Main Text

Scientific output is accelerating faster than the infrastructure supporting its quality. In biomedicine alone, over 1.5 million articles are published annually.^1^ Surveys of the academic workforce consistently show that while scientists spend the vast majority of their time conducting research, they wish they could spend far less time on the administrative and writing tasks required to publish.^2^ However, manuscript preparation is a critical bottleneck; it is usually the part that receives the least attention,^3^despite being the only stage of the research process that is reported to the scientific community.^4^

This discrepancy has consequences. One can present groundbreaking results in a way that easily gets rejected or misunderstood. Content analyses of editorial decisions reveal that “poor presentation”, encompassing poor writing, lack of clarity, and poor elaboration of methods, is one of the most common reasons for both immediate desk rejections and post-peer-review rejections.^5^: If the writing is weak or the formatting is distracting, even the most rigorous methodological findings lose their impact.^6^

To mitigate this, several AI-driven writing tools have emerged, such as Paperpal, Writefull, and Trinka.^7^ These tools provide help and suggestions, yet most of them rely on highly specific, narrow domains. They primarily focus on sentence-level grammar quality, paraphrasing, and basic formatting.^7^ Crucially, none of them look at the project as a whole, nor do they break the evaluation into separable, framework-grounded tasks. Additionally, because these platforms are strictly cloud-based, they introduce data privacy risks that can be bypassed if such a tool were to use local models.

To address these systemic limitations, we introduce Paper Analysis Tool (PAT), a free, multi-agent framework deploying 31 specialized agents powered by locally run small language models (SLMs). PAT can optionally instead use frontier large language models (LLMs) on cloud servers. PAT audits manuscripts across multiple domains. By running open-source entirely on consumer hardware, the system meets the strictest institutional data governance requirements, including HIPAA-compliant workflows while minimizing costs.

PAT operates_through a four-phase, 31-agent architecture that runs entirely on consumer hardware via the Ollama open-source runtime, transmitting zero manuscript content externally.

The pipeline is grounded in three foundational principles. First, determinism: Phase 0 computes six objective text metrics, passive voice ratio, average sentence length, Flesch-Kincaid grade level, long-sentence fraction, hedge-word density, and undefined-acronym count using regular expressions and syllable-counting heuristics, guaranteeing identical output for identical input without any language model involvement Second, framework-grounding: each downstream SLM agent is anchored to an established writing or reporting framework rather than open-ended instruction. Phase 1 deploys sixteen independent agents covering VSNC narrative structure, sentence architecture, voice and tense, conciseness, paragraph quality, acronym hygiene, figures and captions, and reproducibility.^8 9 10^ Phase 2 runs ten whole-document agents’ internal consistency, discussion quality, missing citation detection via PubMed and bioRxiv REST APIs (no API key required), reference verification, and CONSORT/STROBE/TRIPOD+AI^11–13^compliance, each receiving all Phase 1 outputs as prior context. Phase 3 synthesizes every upstream finding into a ranked top 10 action plan with explicit effort-impact scores. Third, transparency: each agent appends a machine-parseable structured footer (severity, 0.0–1.0 quality score, summary, top issues), enabling version-controlled manuscript tracking across revisions and inter-agent agreement quantification via Fleiss’ κ.

**Figure 1.**
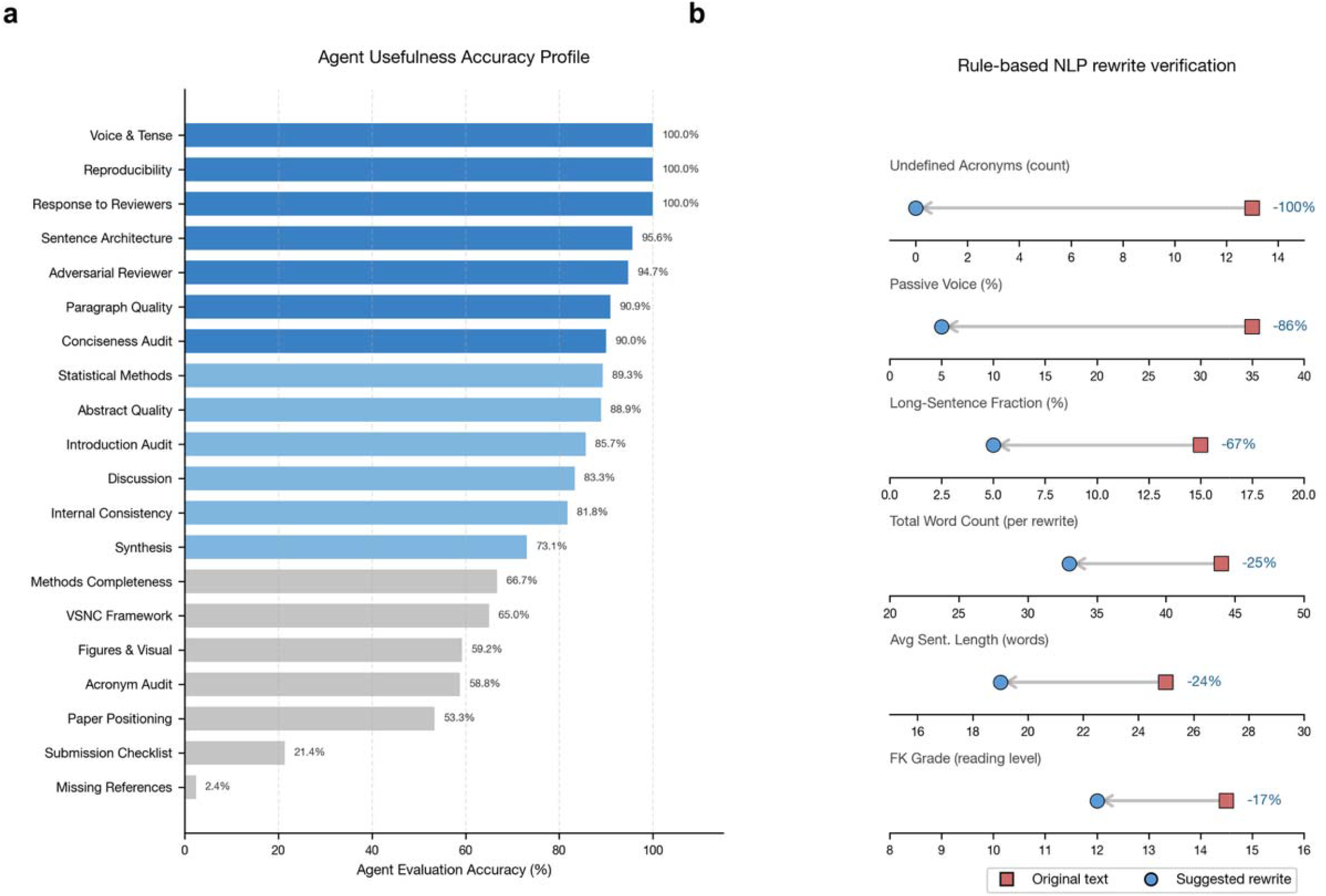
PAT achieves 72.4% suggestion accuracy and reduces passive voice by 86%. a) Agent usefulness accuracy profile across diverse evaluation domains. Two human expert validation demonstrated a 72.4% overall usefulness accuracy, with the multi-agent framework achieving over 95% accuracy in rule-based linguistic domains, including Voice & Tense, Reproducibility, and Sentence Architecture. (b) Rule-based NLP verification of 126 agent-generated original–rewrite passage pairs. Re-evaluation via the deterministic Phase 0 engine confirmed significant, model-unbiased empirical improvements across six core linguistic metrics. The suggested revisions resulted in a 25% reduction in total word count, a sharp decrease in passive voice prevalence from 35% to 5%, a 24% reduction in average sentence length, and a 67% decline in the fraction of overly long sentences. Additionally, the framework drove a 17% improvement in the Flesch–Kincaid reading grade and eliminated undefined clinical acronyms These deterministic textual gains are directionally consistent with the framework’s qualitative usefulness assessments.

**Figure 2.**
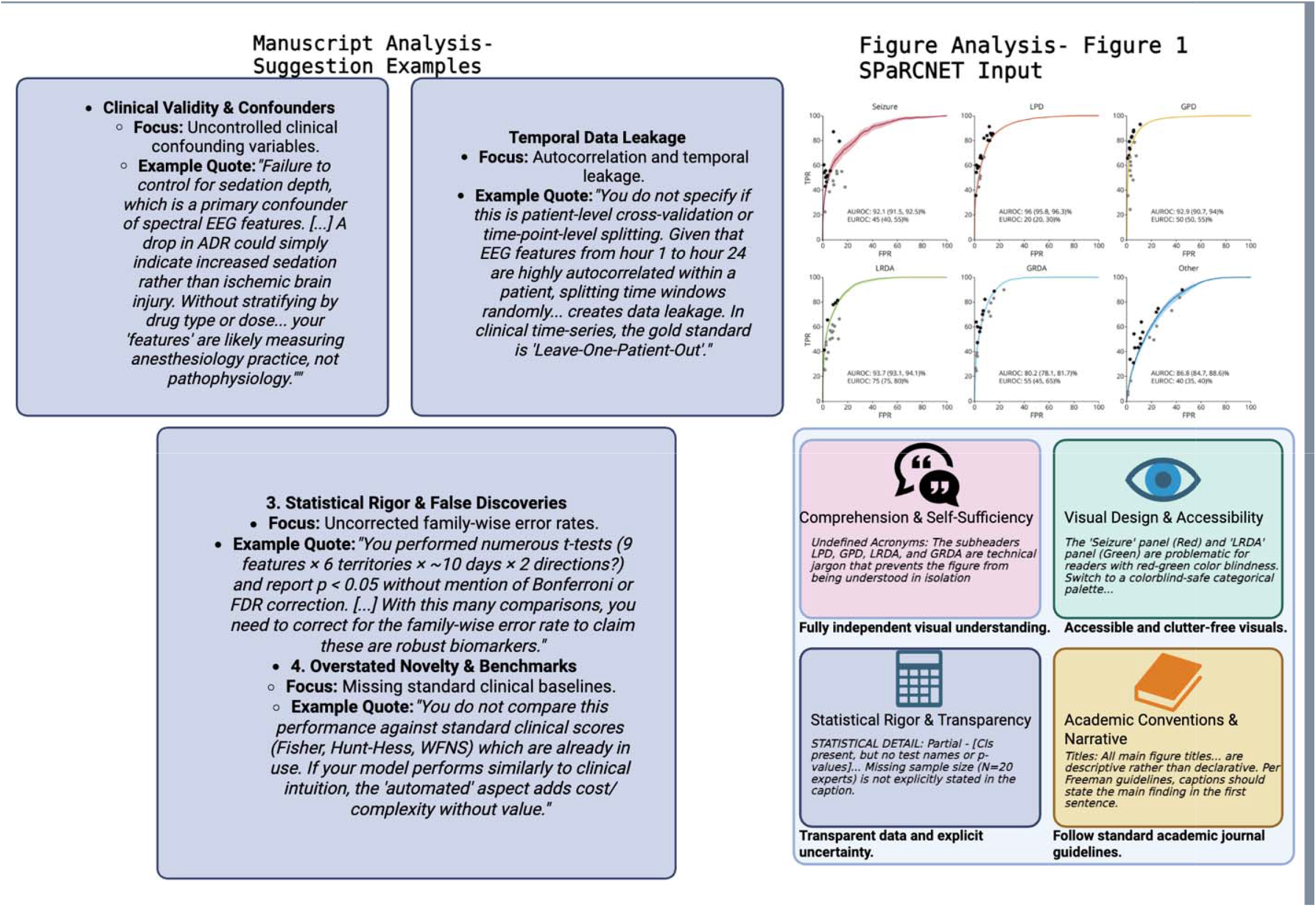
PAT identifies methodological confounders and figure accessibility flaws in representative manuscripts. The agentic framework effectively operated as a high-order peer reviewer, successfully identifying complex, content-dependent methodological vulnerabilities within the text, including uncontrolled clinical confounders and uncorrected family-wise error rates. Concurrently, multimodal vision agents localized accessibility flaws within complex multi-panel diagrams, such as isolating red-green colorblindness hazards and flagged deficits in statistical transparency, including absent sample sizes and missing confidence intervals within foundational figure captions.

PAT showed promise as an AI tool for reviewing manuscripts before submission. Across our three-paper validation corpus, expert human review confirmed that 72.4% of agent-generated suggestions were highly actionable and useful for manuscript improvement, particularly in domains such as Voice & Tense, Reproducibility, and Sentence Architecture. Furthermore, deterministic re-evaluation of 126 suggested rewrite pairs across six rule-based metrics confirmed that these qualitative suggestions translate into measurable, empirical gains including significant reductions in total word count (by 25%), passive voice prevalence (from 35% to 5%), and the elimination of undefined acronyms.

This dual-track validation confirms that the framework successfully targets foundational writing quality bottlenecks, driving measurable and reproducible improvements across multiple linguistic dimensions.

A central crisis in modern scientific literature is the widespread lack of reproducibility and structural rigor.^17^ Many published papers fail to meet fundamental scientific checkpoints, frequently harboring internal inconsistencies, overlapping prior publications, and a profound misalignment between stated methods and supplied code. ^18^,^19^ PAT directly confronts this gap by anchoring its evaluation to established clinical reporting frameworks, such as CONSORT, STROBE, and TRIPOD. ^11–13^ By deploying specialized agents to systematically audit these dimensions, the framework ensures that critical reproducibility checkpoints, which are missing from a vast majority of papers today, are explicitly evaluated before submission. Our validation across published neurological papers confirmed that this systematic pre-submission review drives measurable improvements in structural rigor and reporting compliance

The true impact of this framework lies in its accessibility and its potential to democratize high-quality scientific writing.^13,14^ Commercial AI writing assistants and professional editorial services involve ongoing subscription fees that may be prohibitively expensive for many researchers, particularly those outside well-funded institutions. By removing the reliance on proprietary cloud services, PAT provides a secure, free, and opensource alternative. Any researcher with a standard laptop with 16 GB of VRAM can deploy the system (16 GB VRAM using 4-bit quantized models, or 64 GB+ RAM for full bf16 inference). This removes financial barriers, extending access to sophisticated, multi-modal editing tools to the global research workforce.

Furthermore, the open-source nature of the framework ensures profound adaptability. Because the system features a modular, model-agnostic design, researchers are not locked into a rigid, proprietary ecosystem. Any SLM/LLM can replace the default configuration, allowing the scientific community to freely adjust the model code and swap reasoning engines as newer, more capable open-weight models (or cloud-based models) are released. This flexibility empowers individual laboratories to tailor the agents to their specific operational needs, fine-tune the system for niche journal guidelines, and adjust the framework exactly the way they see fit.

Despite its robust performance, both the framework and our evaluation study possess several notable limitations. Because the system inherently relies on an LLM-as-a-judge paradigm,^22^ it remains susceptible to the standard pitfalls of generative models, including occasional hallucinations, misinterpretations of complex scientific logic, and a lack of absolute precision. Furthermore, while the initial text metrics (Phase 0) are deterministic, the downstream agents possess inherent stochasticity; therefore, executing the pipeline multiple times on the same manuscript may yield slightly different qualitative suggestions across runs. Orchestrating a comprehensive team of 31 specialized agents also introduces significant computational overhead, meaning the complete end-to-end evaluation process may take a couple of hours to execute on local hardware. Regarding the study design, we validated the framework on a limited sample of three previously published papers originating exclusively from our own laboratory, all narrowly focused within the clinical neurology domain. Additionally, expert validation was performed by coauthors involved in tool development (R.B., A.G.), which may introduce confirmation bias; future studies should incorporate blinded evaluation by independent external reviewers. These factors may restrict the immediate generalizability of our findings to other scientific disciplines or formatting styles.

In conclusion, PAT proves that high-quality, framework-grounded manuscript auditing can be achieved locally, openly, and freely, paving the way for more reproducible, rigorous, and accessible scientific literature.

## Methods

### Experimental protocol and agentic pipelines

PAT is a multi-agent framework built on the Ollama open-source runtime (v0.19) for local, privacy-preserving manuscript audit. The pipeline was executed on consumer-grade hardware (128 GB unified RAM) using the Qwen 3.5 27B (bf16) model as the default reasoning engine. The architecture consists of 31 specialized SLM agents deployed in four sequential phases:

*Phase 0*(Deterministic Metrics): An initial text-analysis engine computes six objective metrics without language model involvement: (1) passive voice ratio; (2) average sentence length; (3) Flesch-Kincaid grade level; (4) long-sentence fraction; (5) hedge-word density; and (6) undefined-acronym count. This phase uses standard NLP techniques with deterministic outputs, without any language model involvement

*Phases 1–3* (LLM-as-Judge Pipeline): A panel of 22 text-based agents (Appendix) performs targeted reviews grounded in established writing and reporting frameworks.

*Phase 1 (16 agents, see Appendix)* audits domain-specific dimensions including the VSNC framework (Vision/Steps/News/Contributions), sentence architecture (Gopen &Swan), and reproducibility (methods-code alignment).^8 9 10^

*Phase 2 (10 agents, see Appendix)* evaluates whole-document properties such as internal consistency, “Adversarial Reviewer #2” critique, and compliance with clinical reporting guidelines (CONSORT/STROBE/TRIPOD).

*Phase 3 (4 agents, see Appendix)* synthesizes findings into a ranked action plan, a submission checklist, and a structured Response to Reviewers. A companion vision-capable agentic workflow applies 8 additional agents to audit figure story, composition, and statistical integrity when images are provided. Every agent outputs a machine-parseable structured footer containing a 0.0–1.0 quality score.

## Data availability

Validation papers are publicly available at their respective DOIs. Baseline manuscript texts, full PAT audit reports, and Phase 0 metric extracts are deposited in the project repository at https://github.com/bdsp-core/PAT-PaperAssessmentTool.

## Code availability

PAT source code is available at https://github.com/bdsp-core/PAT-PaperAssessmentTool (MIT Licence). All models download automatically via Ollama on the first run.

## Acknowledgment

Prior to submission, the paper was evaluated using the PAT framework and revised based on its recommendations.

